# Mendelian randomization analysis identified potential genes pleiotropically associated with polycystic ovary syndrome

**DOI:** 10.1101/2021.06.29.21259512

**Authors:** Qian Sun, Gao Yuan, Jingyun Yang, Jiayi Lu, Wen Feng, Wen Yang

**Affiliations:** Department of Gynecology, The First Affiliated Hospital of Kangda College of Nanjing Medical University, Lianyungang, Jiangsu, China; Rush Alzheimer’s Disease Center, Rush University Medical Center, Chicago, IL, USA; Department of Neurological Sciences, Rush University Medical Center, Chicago, IL, USA; Department of Finance, School of Economics, Shanghai University, Shanghai, China

**Keywords:** polycystic ovary syndrome, pleotropic association, expression quantitative trait loci, summary Mendelian randomization

## Abstract

**Research question:** Polycystic ovary syndrome (PCOS) is a common endocrine disorder with unclear etiology. Are there any genes that are pleiotropically or potentially causally associated with PCOS?

**Design:** We applied the summary data-based Mendelian randomization (SMR) method integrating genome-wide association study (GWAS) for PCOS and expression quantitative trait loci (eQTL) data to identify genes that were pleiotropically associated with PCOS. We performed separate SMR analysis using eQTL data in the ovary and whole blood.

**Results:** Although no genes showed significant pleiotropic association with PCOS after correction for multiple testing, some of the genes exhibited suggestive significance. *RPS26* showed the strongest suggestive pleiotropic association with PCOS in both SMR analyses (β[SE]=0.10[0.03], P_SMR_=1.72×10^−4^ for ovary; β[SE]=0.11[0.03], P_SMR_=1.40×10^−4^ for whole blood). *PM20D1* showed the second strongest suggestive pleiotropic association with PCOS in the SMR analysis using eQTL data for the whole blood, and was also among the top ten hit genes in the SMR analysis using eQTL data for the ovary. Two other genes, including *CTC-457L16*.*2* and *NEIL2*, were among the top ten hit genes in both SMR analyses.

**Conclusion:** We identified multiple genes that were potentially involved in the pathogenesis of PCOS. Our findings provided helpful leads to a better understanding of the mechanisms underlying PCOS, and revealed potential therapeutic targets for the effective treatment of PCOS.

**KEY MESSAGE:** Polycystic ovary syndrome (PCOS) is a common endocrine disorder, and its etiology can be multifaceted. We found that multiple genes were potentially involved in the pathogenesis of PCOS. The findings revealed the genetic mechanisms underlying PCOS and potential therapeutic targets for the effective treatment of PCOS.

## INTRODUCTION

Polycystic ovary syndrome (PCOS) is a common endocrine disorder that often occurs in women of childbearing age, characterized by clinical or biochemical manifestations of excessive androgen, persistent anovulation, and polycystic ovarian changes and often accompanied by insulin resistance (IR) and obesity (Bellver et al., 2018). As a highly prevalent endocrine disorder, PCOS affects 5-13% of reproductive-aged women and more than 10% of adolescents (Azziz et al., 2016; Naz et al., 2019; Rotterdam, 2004b; Teede et al., 2010). PCOS is associated with a variety of consequences, such as reproductive issues, metabolic abnormalities, and psychological disorders (Legro et al., 2013). Furthermore, more than half of the PCOS patients suffer from infertility(Joham et al., 2015), and infertile women and adolescent girls with PCOS were reported to have reduced quality of life (QoL) (Barnard et al., 2007; Naumova et al., 2020; Wilson and Pena, 2020).

PCOS is a multifaceted disease, and previous studies have found its association with genetic factors (Zhao et al., 2016), environmental factors such as exposure to Bisphenol A (BPA) (Kandaraki et al., 2011), and life style factors such as smoking (Zhang et al., 2020). However, the exact etiology of PCOS remains to be elucidated. More studies are needed to further explore the pathological mechanisms underlying PCOS to facilitate diagnosis and treatment of this common and jeopardizing disease.

Genetics play an important role in the pathogenesis of PCOS. Previous twin studies demonstrated that genetics explained more than 70% of PCOS pathogenesis (Vink et al., 2006). Multiple genome-wide association studies (GWAS) conducted on Han Chinese, Korean, and European populations have identified multiple genetic variants in association with PCOS, such as single-nucleotide polymorphisms in/near *FSHR, THADA* (Z. J. Chen et al., 2011),*YAP1, HMGA2* (Shi et al., 2012), *KHDRBS3* (Lee et al., 2015), *FSHB, GATA4*/*NEIL2* (Hayes et al., 2015), *ERBB4*/*HER4* and *RAD50* (F. R. Day et al., 2015).

However, biological interpretation of the identified genetic variants in the etiology of PCOS remains largely unclear. It is likely that the genetic variants identified in GWAS could exert their effects on diseases/disorders via gene expression because many of them are located in non-coding regions (Visscher et al., 2012). Therefore, exploring the relationship between genetic variation and gene expression can help better understand the regulatory pathways underlying the pathogenesis of PCOS.

Mendelian randomization (MR) is a method for exploring potentially causal association between an exposure and an outcome by using genetic variants as the instrumental variables (IVs) for exposure (Davey Smith and Hemani, 2014). Compared with traditional statistical methods used in the association studies, MR can reduce confounding and reverse causation and is becoming increasingly popular in the exploration of etiological mechanisms (Burgess et al., 2015; Thanassoulis and O’Donnell, 2009). A novel analytical framework through a summary data-based MR (SMR) approach integrating cis-expression quantitative trait loci (cis-eQTL) or cis-DNA methylation QTL (cis-mQTL) and GWAS data was recently proposed (Zhu et al., 2016), and has been employed in identifying gene expressions or DNA methylation loci that are pleiotropically or potentially causally associated with various phenotypes, such as severity of COVID-19, major depression and Alzheimer’s disease (D. Liu et al., 2020; Pavlides et al., 2016; Wray et al., 2018), implying that it is a promising tool to explore genes that are pleiotropically associated with complex traits.

In this paper, we applied the SMR method integrating summarized GWAS data for PCOS and cis-eQTL data to prioritize genes that are pleiotropically/potentially causally associated with PCOS.

## MATERIALS AND METHODS

### Data sources

#### eQTL data

In the SMR analysis, cis-eQTL genetic variants were used as the instrumental variables (IVs) for gene expression. We performed SMR analysis using eQTL data in ovary and blood from the V7 release of the GTEx summarized data. The summarized data included 85 participants for ovary and 338 participants for blood (Consortium et al., 2017). The eQTL data can be downloaded at https://cnsgenomics.com/data/SMR/#eQTLsummarydata.

#### GWAS data for PCOS

The GWAS summarized data for CCT were provided by a recent genome-wide association meta-analysis of PCOS(F. Day et al., 2018). The results were based on meta-analyses of seven cohorts of European descent, including a total of 10,074 PCOS cases and 103,164 controls. Diagnosis of PCOS was based on the NIH (Zawadzki and Dunaif, 1992) or the Rotterdam Criteria (Rotterdam, 2004a), and the self-report-based data from 23andMe were excluded. The GWAS summarized data can be downloaded at https://www.repository.cam.ac.uk/handle/1810/283491.

#### SMR analysis

In the SMR analyses, cis-eQTL was the IV, gene expression was the exposure, and PCOS was the outcome. The analysis was done using the method as implemented in the software SMR. SMR applies the principles of MR to jointly analyze GWAS and eQTL summary statistics to test for pleotropic association between gene expression and a trait due to a shared and potentially causal variant at a locus. Detailed information regarding the SMR method was reported in a previous publication (Zhu et al., 2016). We also conducted the heterogeneity in dependent instruments (HEIDI) test to evaluate the existence of linkage in the observed association. Rejection of the null hypothesis (i.e., P_HEIDI_<0.05) indicates that the observed association could be due to two distinct genetic variants in high linkage disequilibrium with each other. We adopted the default settings in SMR (e.g., P_eQTL_ <5 × 10^−8^, minor allele frequency [MAF] > 0.01, removing SNPs in very strong linkage disequilibrium [LD, r^2^ > 0.9] with the top associated eQTL, and removing SNPs in low LD or not in LD [r^2^ <0.05] with the top associated eQTL), and used false discovery rate (FDR) to adjust for multiple testing.

Data cleaning and statistical/bioinformatical analysis was performed using R version 4.0.4 (https://www.r-project.org/), PLINK 1.9 (https://www.cog-genomics.org/plink/1.9/) and SMR (https://cnsgenomics.com/software/smr/).

## RESULTS

### Basic information of the summarized data

The GWAS summarized data were based on GWAS meta-analysis of 113,238 subjects (10,074 PCOS cases and 103,164 controls). After checking of allele frequencies among the datasets and LD pruning, there were about 6.4 million eligible SNPs included in the final SMR analysis. The number of participants for GETx eQTL data in ovary is smaller (n=85), compared with that in whole blood (n=385), so is the number of eligible probes (1,530 vs. 4,490). The detailed information was shown in **Table 1**.

**Table 1.** Basic information of the GWAS and eQTL data.

| **Data Source** | **Total number of participants** | **Number of eligible genetic variants or probes** |
| --- | --- | --- |
| **eQTL data** |  |  |
| **Ovary** | 85 | 1,530 |
| **Whole blood** | 338 | 4,490 |
| **GWAS data** | 113,238 | 6,400,755 |

GWAS: genome-wide association studies; QTL, quantitative trait loci

### Pleiotropic association with PCOS

Information of the top ten probes using eQTL data for the ovary and whole blood was presented in **Table 2**. Although no genes showed significant pleiotropic association with PCOS after correction for multiple testing, some of genes exhibited suggestive significance. Specifically, *RPS26* (ENSG00000197728.5) showed the strongest suggestive pleiotropic association with PCOS in both SMR analyses (β[SE]=0.10[0.03], P_SMR_=1.72×10^−4^ for ovary; β[SE]=0.11[0.03], P_SMR_=1.40×10^−4^ for whole blood; **Figure 1**). *PM20D1* (ENSG00000162877.8) showed the second strongest suggestive pleiotropic association with PCOS in the SMR analysis using eQTL data for the whole blood, and it was also among the top ten hit genes in the SMR analysis using eQTL data for the ovary (β[SE]=0.10[0.03], P_SMR_=2.94×10^−3^ for ovary; β[SE]=0.14[0.04], P_SMR_=8.38×10^−4^ for blood; **Figure 2**). Two other genes, including *CTC-457L16*.*2* (ENSG00000262319.1; β[SE]=-0.2[0.06], P_SMR_=1.55×10^−3^ for ovary; β[SE]=-0.31[0.09], P_SMR_=1.07×10^−3^ for blood; **Figure 3**) and *NEIL2* (ENSG00000154328.11; β[SE]=-0.2[0.06], P_SMR_=1.02×10^−3^for ovary; β[SE]=-0.30[0.10], P_SMR_=2.25×10^−3^ for blood; **Figure 4**), were among the top ten hit genes in both SMR analyses.

**Table 2.** The top ten probes identified in the SMR analysis.

| eQTL data | Probe ID | Gene | CHR | Top SNP | P<sub>eQTL</sub> | P<sub>GWAS</sub> | Beta | SE | P<sub>SMR</sub> | P<sub>HEIDI</sub> | N<sub>SNP</sub> |
| --- | --- | --- | --- | --- | --- | --- | --- | --- | --- | --- | --- |
| **Ovary** | ENSG00000197728.5 | *RPS26* | 12 | rs1131017 | 3.89×10<sup>-55</sup> | 1.60×10<sup>-4</sup> | 0.10 | 0.03 | 1.72×10<sup>-4</sup> | 1.87×10<sup>-1</sup> | 20 |
|  | ENSG00000261390.1 | *RP11-345M22.2* | 16 | rs7196490 | 1.02×10<sup>-11</sup> | 1.20×10<sup>-5</sup> | -0.28 | 0.08 | 3.18×10<sup>-4</sup> | 8.51×10<sup>-1</sup> | 20 |
|  | ENSG00000154328.11 | *NEIL2* | 8 | rs804271 | 1.71×10<sup>-14</sup> | 3.70×10<sup>-4</sup> | -0.20 | 0.06 | 1.02×10<sup>-3</sup> | 1.87×10<sup>-1</sup> | 20 |
|  | ENSG00000225706.1 | *RP11-75C9.1* | 9 | rs77301379 | 7.45×10<sup>-20</sup> | 5.30×10<sup>-4</sup> | 0.13 | 0.04 | 1.41×10<sup>-3</sup> | 8.71×10<sup>-1</sup> | 20 |
|  | ENSG00000262319.1 | *CTC-457L16.2* | 17 | rs55885404 | 3.59×10<sup>-9</sup> | 1.80×10<sup>-4</sup> | -0.20 | 0.06 | 1.55×10<sup>-3</sup> | 9.58×10<sup>-1</sup> | 12 |
|  | ENSG00000172000.3 | *ZNF556* | 19 | rs4807349 | 2.34×10<sup>-10</sup> | 5.70×10<sup>-4</sup> | -0.24 | 0.08 | 2.24×10<sup>-3</sup> | 1.84×10<sup>-1</sup> | 20 |
|  | ENSG00000162877.8 | *PM20D1* | 1 | rs823080 | 3.53×10<sup>-22</sup> | 8.50×10<sup>-4</sup> | 0.10 | 0.03 | 2.94×10<sup>-3</sup> | 9.31×10<sup>-1</sup> | 20 |
|  | ENSG00000186300.7 | *ZNF555* | 19 | rs917653 | 3.31×10<sup>-12</sup> | 8.30×10<sup>-4</sup> | -0.15 | 0.05 | 3.18×10<sup>-3</sup> | 8.66×10<sup>-1</sup> | 17 |
|  | ENSG00000165623.5 | *UCMA* | 10 | rs78415540 | 1.12×10<sup>-20</sup> | 3.80×10<sup>-3</sup> | -0.09 | 0.03 | 3.69×10<sup>-3</sup> | 1.46×10<sup>-1</sup> | 20 |
|  | ENSG00000100211.6 | *CBY1* | 22 | rs62229988 | 3.09×10<sup>-13</sup> | 2.50×10<sup>-3</sup> | 0.21 | 0.07 | 3.90×10<sup>-3</sup> | 6.00×10<sup>-1</sup> | 20 |
| **Whole blood** | ENSG00000197728.5 | *RPS26* | 12 | rs1131017 | 5.82×10<sup>-101</sup> | 1.60×10<sup>-4</sup> | 0.11 | 0.03 | 1.40×10<sup>-4</sup> | 3.60×10<sup>-1</sup> | 20 |
|  | ENSG00000162877.8 | *PM20D1* | 1 | rs1772143 | 3.17×10<sup>-45</sup> | 5.20×10<sup>-4</sup> | 0.14 | 0.04 | 8.38×10<sup>-4</sup> | 8.78×10<sup>-1</sup> | 20 |
|  | ENSG00000262319.1 | *CTC-457L16.2* | 17 | rs55885404 | 2.28×10<sup>-11</sup> | 1.80×10<sup>-4</sup> | -0.31 | 0.09 | 1.07×10<sup>-3</sup> | 9.96×10<sup>-1</sup> | 16 |
|  | ENSG00000100201.14 | *DDX17* | 22 | rs2267390 | 5.13×10<sup>-18</sup> | 4.90×10<sup>-4</sup> | -0.77 | 0.24 | 1.08×10<sup>-3</sup> | 3.23×10<sup>-2</sup> | 20 |
|  | ENSG00000127483.13 | *HP1BP3* | 1 | rs9426742 | 2.54×10<sup>-16</sup> | 6.90×10<sup>-4</sup> | 0.60 | 0.19 | 1.53×10<sup>-3</sup> | 9.17×10<sup>-1</sup> | 20 |
|  | ENSG00000242550.1 | *SERPINB10* | 18 | rs9955526 | 4.76×10<sup>-37</sup> | 1.00×10<sup>-3</sup> | 0.22 | 0.07 | 1.64×10<sup>-3</sup> | 1.11×10<sup>-1</sup> | 20 |
|  | ENSG00000128482.11 | *RNF112* | 17 | rs7502682 | 2.34×10<sup>-20</sup> | 9.90×10<sup>-4</sup> | 0.23 | 0.07 | 1.71×10<sup>-3</sup> | 2.58×10<sup>-1</sup> | 20 |
|  | ENSG00000254415.3 | *SIGLEC14* | 19 | rs872629 | 2.70×10<sup>-30</sup> | 1.20×10<sup>-3</sup> | 0.20 | 0.07 | 1.97×10<sup>-3</sup> | 2.71×10<sup>-1</sup> | 20 |
|  | ENSG00000154328.11 | *NEIL2* | 8 | rs804270 | 2.11×10<sup>-14</sup> | 1.00×10<sup>-3</sup> | -0.30 | 0.10 | 2.25×10<sup>-3</sup> | 1.33×10<sup>-1</sup> | 20 |
|  | ENSG00000105501.7 | *SIGLEC5* | 19 | rs872629 | 7.55×10<sup>-14</sup> | 1.20×10<sup>-3</sup> | 0.52 | 0.18 | 3.15×10<sup>-3</sup> | 1.83×10<sup>-1</sup> | 18 |

The GWAS summarized data were provided by the study of Shungin et al. and can be downloaded at https://www.repository.cam.ac.uk/handle/1810/283491. The GTEx eQTL data can be downloaded at https://cnsgenomics.com/data/SMR/#eQTLsummarydata.
P<sub>eQTL</sub> is the P-value of the top associated cis-eQTL in the eQTL analysis, and P<sub>GWAS</sub> is the P-value for the top associated cis-eQTL in the GWAS analysis. Beta is the estimated effect size in SMR analysis, SE is the corresponding standard error, P<sub>SMR</sub> is the P-value for SMR analysis, P<sub>HEIDI</sub> is
the P-value for the HEIDI test and $N_{\text{snp}}$ is the number of SNPs involved in the HEIDI test. FDR was calculated at $P=10^{-3}$ threshold.
**Bold font** means statistical significance after correction for multiple testing using FDR.
CHR, chromosome; HEIDI, heterogeneity in dependent instruments; SNP, single-nucleotide polymorphism; SMR, summary data-based Mendelian randomization; QTL, quantitative trait loci; FDR, false discovery rate; GWAS, genome-wide association studies

**Figure 1.**
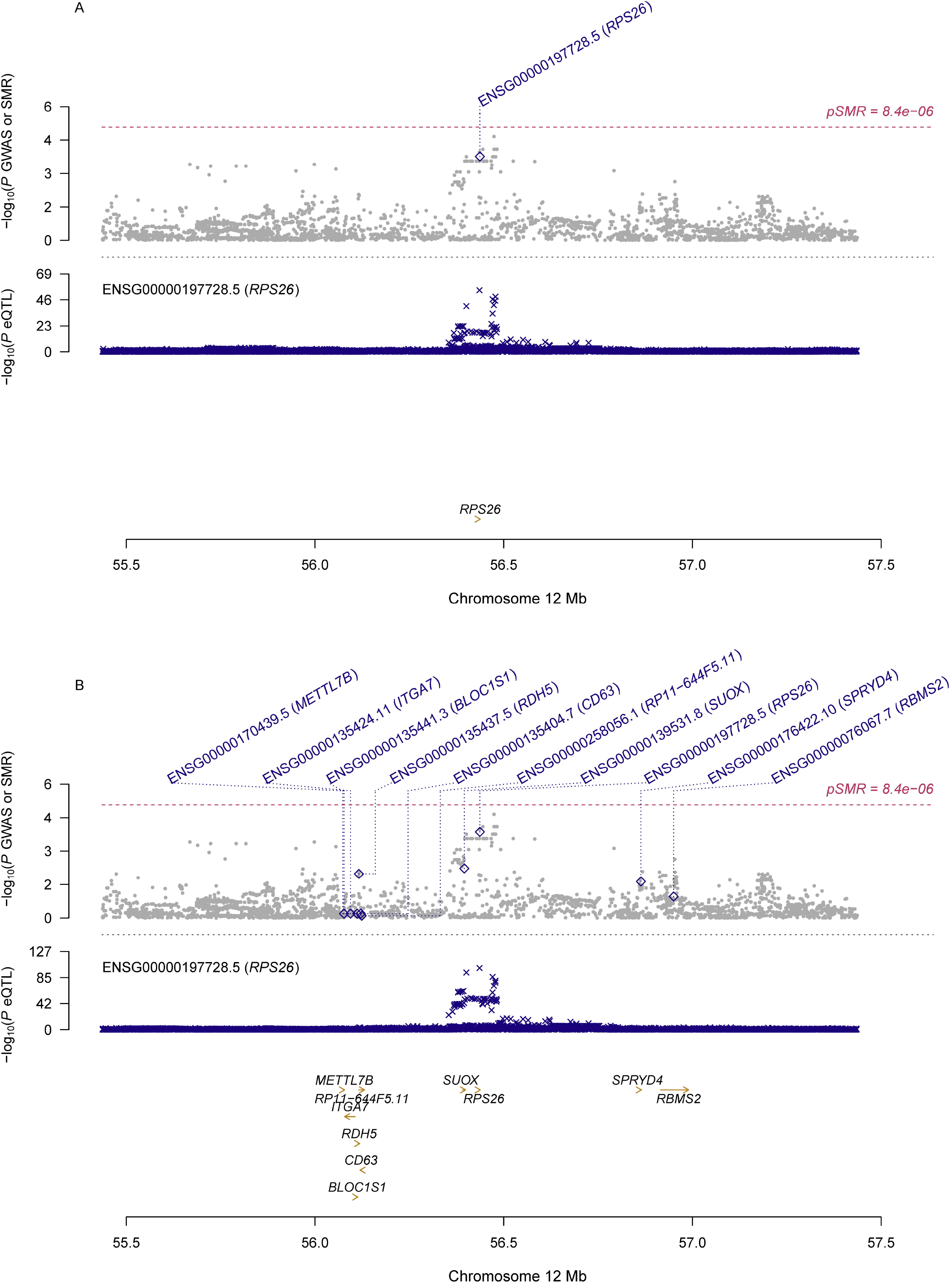
Pleiotropic association of *RPS26* with PCOS. A) SMR analysis results using eQTL data for ovary; B) SMR analysis results using eQTL data for whole blood Top plot, grey dots represent the -log_10_(*P* values) for SNPs from the GWAS of PCOS, with solid rhombuses indicating that the probes pass HEIDI test. Middle plot, eQTL results. Bottom plot, location of genes tagged by the probes. GWAS, genome-wide association studies; SMR, summary data-based Mendelian randomization; HEIDI, heterogeneity in dependent instruments; eQTL, expression quantitative trait loci; PCOS, polycystic ovary syndrome

**Figure 2.**
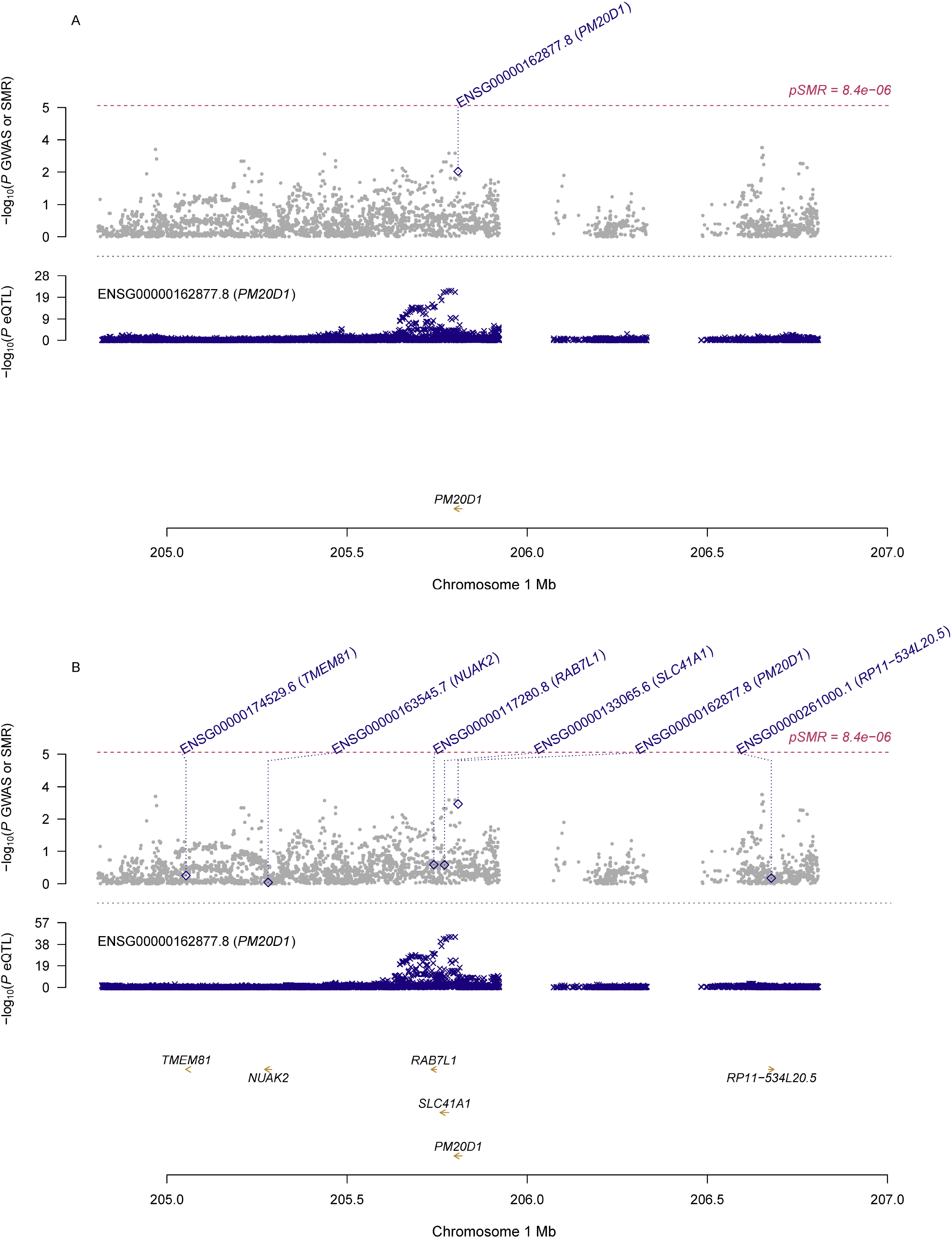
Pleiotropic association of *PM20D1* with PCOS. A) SMR analysis results using eQTL data for ovary; B) SMR analysis results using eQTL data for whole blood Top plot, grey dots represent the -log_10_(*P* values) for SNPs from the GWAS of PCOS, with solid rhombuses indicating that the probes pass HEIDI test. Middle plot, eQTL results. Bottom plot, location of genes tagged by the probes. GWAS, genome-wide association studies; SMR, summary data-based Mendelian randomization; HEIDI, heterogeneity in dependent instruments; eQTL, expression quantitative trait loci; PCOS, polycystic ovary syndrome

**Figure 3.**
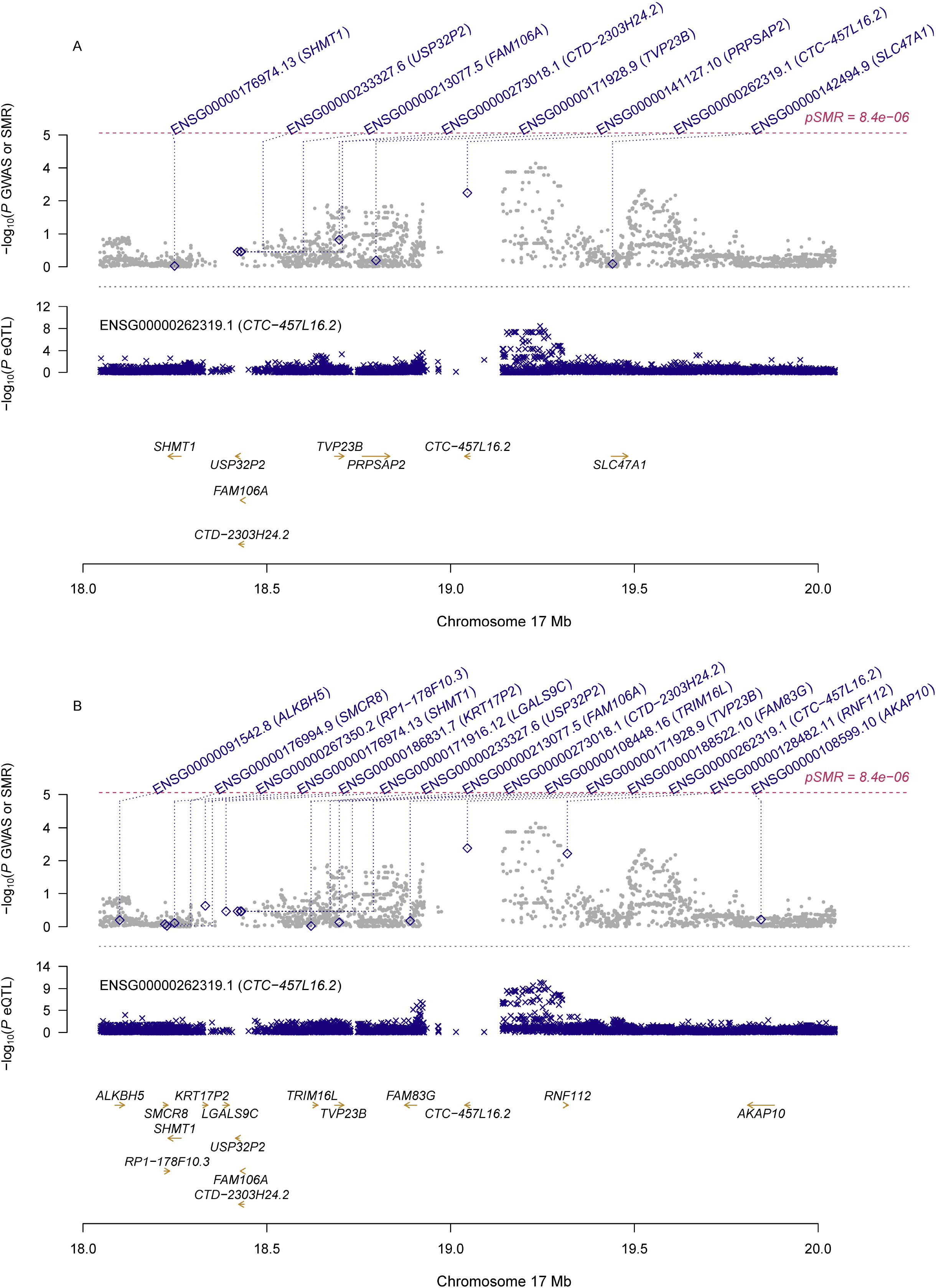
Pleiotropic association of *CTC-457L16*.*2* with PCOS. A) SMR analysis results using eQTL data for ovary; B) SMR analysis results using eQTL data for whole blood Top plot, grey dots represent the -log_10_(*P* values) for SNPs from the GWAS of PCOS, with solid rhombuses indicating that the probes pass HEIDI test. Middle plot, eQTL results. Bottom plot, location of genes tagged by the probes. GWAS, genome-wide association studies; SMR, summary data-based Mendelian randomization; HEIDI, heterogeneity in dependent instruments; eQTL, expression quantitative trait loci; PCOS, polycystic ovary syndrome

**Figure 4.**
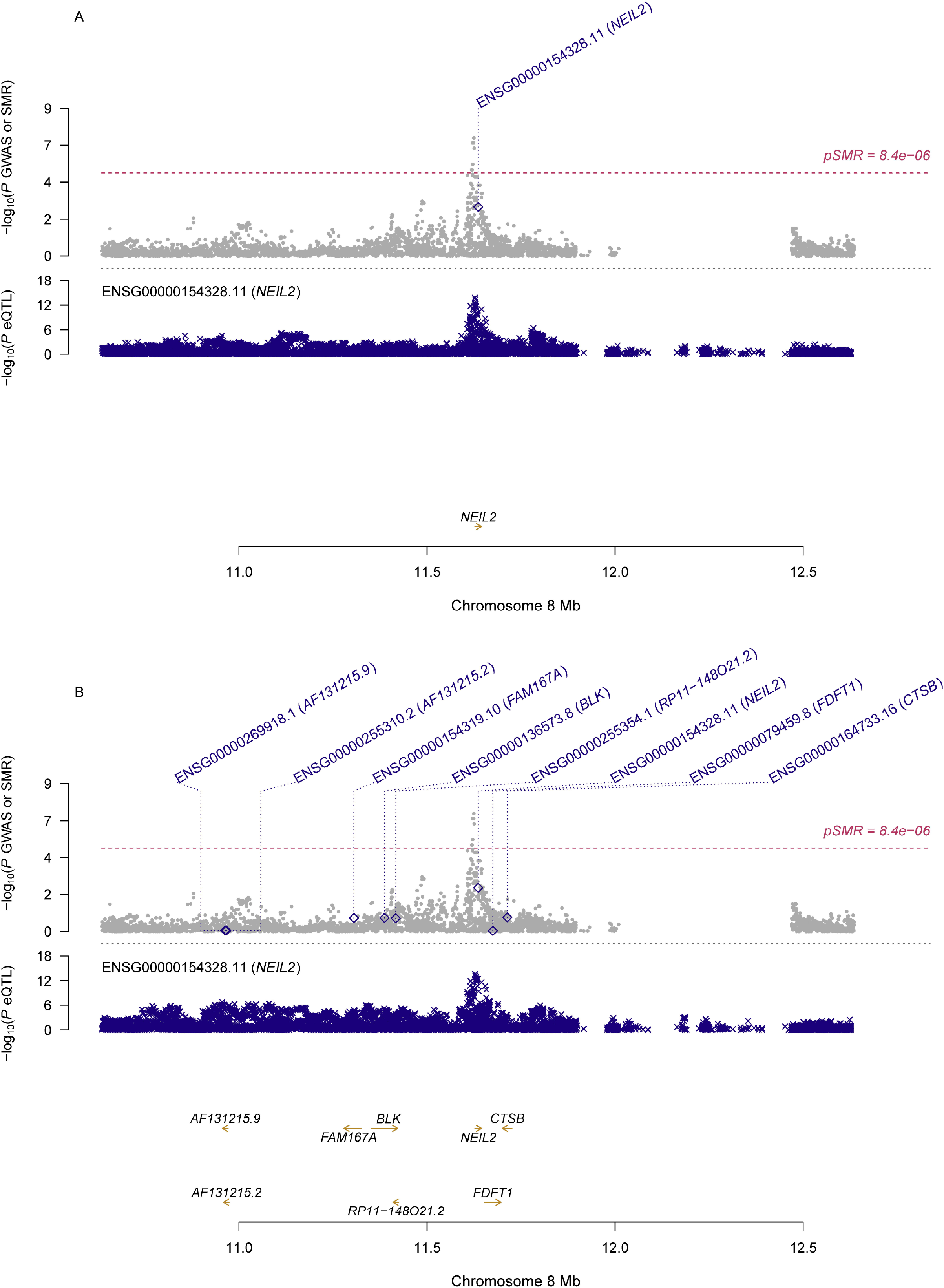
Pleiotropic association of *NEIL2* with PCOS. A) SMR analysis results using eQTL data for ovary; B) SMR analysis results using eQTL data for whole blood Top plot, grey dots represent the -log_10_(*P* values) for SNPs from the GWAS of PCOS, with solid rhombuses indicating that the probes pass HEIDI test. Middle plot, eQTL results. Bottom plot, location of genes tagged by the probes. GWAS, genome-wide association studies; SMR, summary data-based Mendelian randomization; HEIDI, heterogeneity in dependent instruments; eQTL, expression quantitative trait loci; PCOS, polycystic ovary syndrome

## DISCUSSION

In this study, we integrated GWAS summarized data for PCOS and eQTL data in the SMR analysis to explore genes that were pleiotropically or potentially causal associated with PCOS. We found that multiple genes were potentially involved in the pathogenesis of PCOS. To the best of our knowledge, this is the first study to explore genes in pleiotropic association PCOS through a Mendelian randomization approach. Our findings provided helpful leads to a better understanding of the mechanisms underlying PCOS, and revealed potential therapeutic targets for the effective treatment of PCOS.

In our study, *RPS26* (Ribosomal Protein S26) showed the strongest suggestive pleiotropic association with PCOS in the SMR analyses using both ovary and blood eQTL data (**Table 2**). *RPS26*, located on 12q13.2, is a gene encoding a ribosomal protein which is a component of the 40S subunit and belongs to the S26E family of ribosomal proteins (Filipenko et al., 1998). A recent research found that RPS26 critically regulated T-cell survival in a p53-dependent manner (C. Chen et al., 2021). Knockout of *RPS26* in mouse oocytes led to retarded follicle development from pre-antral follicles to antral follicles, and arrested chromatin configurations of the oocytes (X. M. Liu et al., 2018). An earlier study using human ovary cDNA library found that RPS26 was downregulated in PCOS ovary, compared with the normal human ovary (Diao et al., 2004). A recent study examined genes in PCOS-associated regions using a Bayesian colocalization approach (Coloc) and found seven genes, including *RPS26*, that harbored potential causal variants accounting for approximately 30% of known PCOS signals (Censin et al., 2021). These findings, together with ours, suggested that *RPS26* might play a critical role in the etiology of PCOS, and highlighted the potential of this gene as a promising target for the prevention and treatment of PCOS.

*PM20D1* (Peptidase M20 Domain Containing 1) showed the second strongest suggestive pleiotropic association with PCOS in the SMR analysis using blood eQTL data, and was also among the top hit genes in the SMR analysis using ovary eQTL data (**Table 2**). *PM20D1*, located on 1q32.1, is a gene encoding a bidirectional enzyme capable of catalyzing both the condensation of fatty acids and amino acids to generate N-acyl amino acids and also the reverse hydrolytic reaction, thereby regulating energy homeostasis (Long et al., 2016). Mice with increased circulating PM20D1 had increased N-acyl amino acids in blood and improved glucose homeostasis, and *PM20D1* knockout in mice resulted in impaired glucose tolerance and insulin resistance (IR)(Long et al., 2018). In human adipocytes, *PM20D1* is one of the most highly up-regulated genes by the antidiabetic thiazolidinedione drug rosiglitazone, suggesting a potential role of this enzyme and/or N-fatty acyl amino acids in obesity and diabetes (Benson et al., 2019). In addition, *PM20D1* has been found to be associated with various diseases such as Parkinson’s disease (Pihlstrom et al., 2015) and Alzheimer’s disease (Sanchez-Mut et al., 2018). To date, research is scarce on the association of *PM20D1* with PCOS. Given that PCOS is a metabolic disorder that is closely related with obesity, IR, type 2 diabetes (T2D) and metabolic syndrome (Condorelli et al., 2017), it is highly likely that *PM20D1* could be involved in the etiology of PCOS, and more studies are needed to explore the exact roles that *PM20D1* plays in the pathogenesis of PCOS.

*NEIL2* (Nei Like DNA Glycosylase 2) showed the third strongest suggestive pleiotropic association with PCOS in the SMR analysis using ovary eQTL data, and was also among the top hit genes in the SMR analysis using whole blood eQTL data (**Table 2**). *NEIL2*, located on 8p23.1, is a gene encoding a member of the Fpg/Nei family of DNA glycosylases. The encoded enzyme is primarily involved in DNA repair by cleaving oxidatively damaged bases and introducing a DNA strand break via its abasic site lyase activity (Dou et al., 2003; Hazra et al., 2002). The recent meta-analysis of GWAS on PCOS, on which our SMR analysis was based, found that the genetic variant rs804279 in *GATA4*/*NEIL2* showed significant association with PCOS (OR=1.14, 95% CI: 1.10-1.18; P=3.76×10^−12^); however, significant heterogeneity was observed across the different studies (F. Day et al., 2018). This genetic variant also showed significant association with polycystic ovarian morphology and ovulatory dysfunction (F. Day et al., 2018). Another genetic variant rs8191514 in *NEIL2* was predicted to generate a binding site for twenty transcription factors, and it was in linkage disequilibrium with the PCOS-identified genetic variant rs804279 (R^2^= 0.4, D □ =0.97; P<0.0001) (Prabhu et al., 2021). Moreover, 8p23.1, where the *NEIL2* is located, is also the region of *GATA4* (GATA Binding Protein 4), and knock-out of *GATA4* led to abnormal responses to exogenous gonadotropins and impaired fertility in mice (LaVoie, 2014). It also encompasses the promoter region of *FDFT1* (Farnesyl-Diphosphate Farnesyltransferase 1), a gene encoding farnesyl-diphosphate farnesyl transferase that is involved in cholesterol biosynthesis pathway (Ha and Lee, 2020), thereby influencing testosterone biosynthesis. These findings, together with ours, highlight the importance of this region in association with PCOS.

Our study has some limitations. The number of probes used in our SMR analyses was limited, and the sample size in the eQTL analysis was limited, especially for the eQTL data in ovary, which may lead to reduced power in the eQTL analysis. Consequently, we may have missed some important genes implicated in PCOS. Future SMR studies with larger samples for the eQTL analysis are warranted to identify additional genes underlying the pathogenesis of PCOS. We only performed SMR analyses in participants of European ethnicity, and future studies are needed to explore whether our findings can be generalized to other ethnicities. Moreover, we only explored gene expression probes in association with PCOS, and it is possible that genetic variants exert their effect on PCOS through other epigenetic mechanisms, such as DNA methylation. More studies integrating multi-omics data are needed to more systematically explore the complex mechanisms underpinning PCOS.

In summary, our SMR study integrating GWAS of PCOS and eQTL data revealed multiple genes were potentially involved in the pathogenesis of PCOS. Some of the genes were reported to be involved in the regulation of T-cell survival, energy homeostasis, and DNA repair. More studies are needed to examine the exact functions of these genes in the etiology of PCOS and to explore additional genes implicated in the pathogenesis of PCOS.

## Data Availability

All data generated or analyzed during this study are publicly available as specified in the methods section of this paper. The eQTL data can be downloaded at https://cnsgenomics.com/data/SMR/#eQTLsummarydata, and the GWAS summarized data can be downloaded at https://www.repository.cam.ac.uk/handle/1810/283491.

## ABBREVIATIONS

BPA: Bisphenol A;
eQTL: expression quantitative trait loci;
GWAS: genome-wide association study;
HEIDI: heterogeneity in dependent instruments;
IR: insulin resistance;
MR: mendelian randomization;
PCOS: polycystic ovary syndrome;
QoL: quality of life;
SMR: summary data-based Mendelian randomization

## ACKNOWLEDGEMENTS

This work was supported by Lianyungang Health and Family Planning Science and Technology Project (No. 201706). Dr. Jingyun Yang’s research was supported by NIH/NIA grants [P30AG10161, R01AG15819, R01AG17917, R01AG36042, U01AG61356 and 1RF1AG064312-01].

## COMPETING INTERESTS

No potential conflicts of interest were disclosed by the authors.

## AUTHORS’ CONTRIBUTIONS

QS, JY, WF and WY designed and registered the study. DL, JL and JY analyzed data and performed data interpretation. QS, YG, DL and JY wrote the initial draft and JY and WY contributed writing to subsequent versions of the manuscript. All authors reviewed the study findings and read and approved the final version before submission.

## Notes

### Competing Interest Statement

The authors have declared no competing interest.

### Clinical Protocols

https://www.r-project.org/

https://www.cog-genomics.org/plink/1.9/

https://cnsgenomics.com/software/smr/

### Author Declarations

This work was approved by the Ethics committee of the First Affiliated Hospital of Kangda College of Nanjing Medical University.

